# Multimodal artificial intelligence for personalized hepatocellular carcinoma treatment strategy selection

**DOI:** 10.64898/2026.08.21.26361067

**Authors:** Wei Feng, Suyi Liu, Zhiyun Yang, Yiru Tao, Xiangqian Gu, Wei Jin

## Abstract

**Background:** Hepatocellular carcinoma (HCC) treatment selection demands nuanced integration of heterogeneous patient data, yet prevailing predictive models rely on restricted data modalities and oversimplified therapeutic frameworks, compromising clinical translation.

**Objective:** We developed and validated a multimodal artificial intelligence framework to guide optimal treatment strategy selection across the full spectrum of HCC interventions.

**Methods:** This retrospective study comprised 1,043 HCC patients (development cohort, January 2017–December 2023) and 55 external validation patients (2023) from Wuxi People’s Hospital. We engineered Embedding-Augmented Extra Trees (ET-Emb), a novel model fusing structured clinical variables with contextual text embeddings derived from medical histories and radiology reports. ET-Emb quantifies probabilities for five primary treatments: open/laparoscopic resection, transarterial chemoembolization, radiofrequency ablation (RFA), and chemotherapy. Model performance was rigorously assessed via 10-fold cross-validation and external validation using ROC-AUC and PR-AUC metrics.

**Results:** ET-Emb demonstrated robust performance in the development cohort (ROC-AUC: 0.84 ± 0.04; PR-AUC: 0.55 ± 0.06), significantly outperforming established benchmarks. This generalizability was preserved in external validation (ROC-AUC: 0.77 ± 0.02; PR-AUC: 0.47 ± 0.03). SHAP analysis identified textual clinical narratives and socioeconomic determinants as critical predictive drivers.

**Conclusions:** By unifying structured and unstructured data modalities, ET-Emb delivers accurate, multi-treatment strategy prediction for HCC. Its clinical validity and the demonstrated significance of textual features establish multimodal AI as an essential paradigm for simulating complex oncological decision-making, positioning ET-Emb as a transformative tool for precision HCC management.

## Introduction

Hepatocellular carcinoma (HCC) ranks third globally and second in China in terms of cancer-related mortality, nearly half of all global liver cancer cases occur in China[1]. HCC accounts for 75% to 85% of primary liver cancers. The treatment of HCC is characterized by a multidisciplinary approach involving various therapeutic modalities, including hepatic resection, ablation, transarterial chemoembolization (TACE), radiation therapy, and systemic antitumor therapies[2].

Therapeutic selection for HCC patients requires a integration of tumor staging, baseline hepatic function, and patient performance status[2]. Determinants such as tumor anatomical location, disease distribution, and comorbid conditions influence clinical outcomes. For example, patients with CNLC stage IIa HCC may be eligible for multiple interventions— including surgical resection, TACE, or systemic chemotherapy. However, additional variables like socioeconomic factors, and patient preferences frequently dictate the final treatment decision, making the process highly intricate and often subjective[2]. This inherent complexity underscores a critical need for more objective and comprehensive decision-making tools to ensure patients receive the most appropriate and effective treatment.

Existing studies primarily leverage structured clinical data and employ machine learning models to predict treatment modalities[3,4]. Zhang et al. demonstrated the predictive capability of radiomic features for post-recurrence treatment selection[5]. However, these investigations suffer from two key limitations: insufficient integration of diverse multimodal data and restricted coverage of the full spectrum of available therapeutic options. The reliance on limited data types fails to capture the holistic patient profile that clinicians consider, and the narrow scope of treatment predictions does not reflect the multifactorial nature of real-world HCC management.

To address these critical limitations, our study aims to develop a novel AI model, ET-Emb, that accurately mimics the complex clinical decision-making processes for HCC by comprehensively incorporating pre-treatment multimodal data. Our objective is to integrate demographic characteristics, clinical parameters, imaging biomarkers, and socioeconomic factors to provide optimal therapeutic recommendations. We hypothesize that by leveraging this rich, multimodal dataset, the ET-Emb model will achieve robust predictive performance across various evaluation metrics, thereby offering a more comprehensive and clinically relevant tool for HCC treatment guidance.

## Methods

This retrospective study adhered to the Declaration of Helsinki and was approved by the Ethics Review Board of Wuxi People’s Hospital, Nanjing Medical University (KY24160). Written informed consent was waived due to the retrospective nature of the study and full anonymization of data via hospital ID coding.

### Development cohort

Patients with HCC (HCC; ICD-10 code C22) treated at Wuxi People’s Hospital, Nanjing Medical University, between January 2017 and December 2024 were included. A total of 1894 patients were initially identified through the hospital’s medical record management system. The selection process (Fig 1) involved the following exclusion criteria: (1) exclusion of 47 patients with a prior history of surgery; (2) exclusion of 314 patients with metastatic liver cancer or secondary hepatic malignancies; (3) exclusion of 71 patients with concomitant cholangiocarcinoma or other non-hepatocellular carcinoma liver tumors; (4) exclusion of 7 patients who underwent liver transplantation; and (5) exclusion of 412 patients who did not receive imaging assessments via ultrasound, computed tomography (CT), or magnetic resonance imaging (MRI). A final cohort of 1043 patients was included in the study.

**Fig 1.**
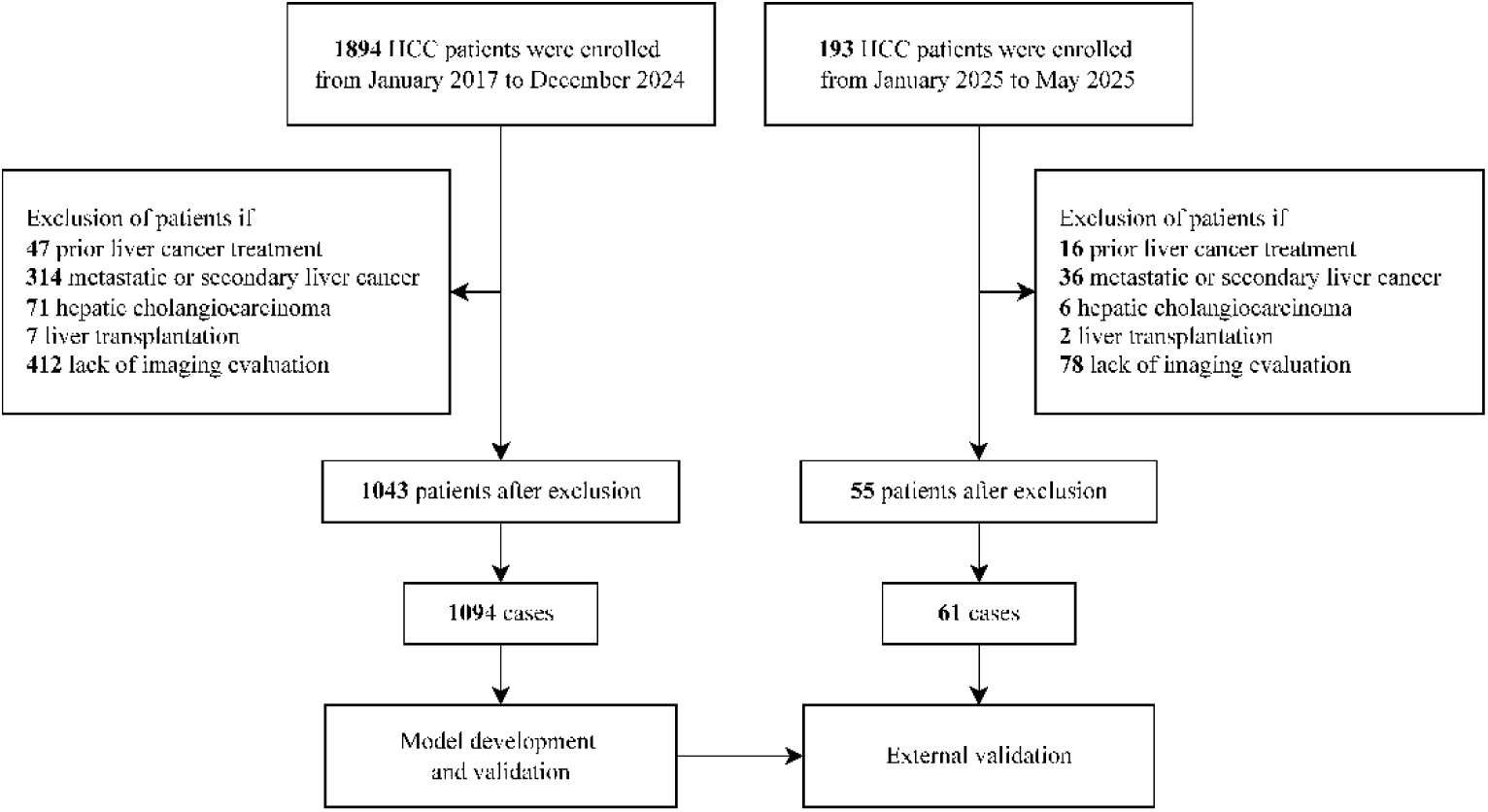
Flowchart of the construction of the development cohort and the external validation cohort.

### External validation cohort

The external validation cohort consisted of 55 patients with HCC admitted to Wuxi People’s Hospital from January 1, 2025, to May 31, 2025, after applying the same exclusion criteria used for the development cohort.

### Variables

#### Co-variates

To capture a comprehensive representation of patient characteristics and disease status, a total of 100 clinical features were extracted from the inpatient records of 1,043 patients, covering four categories: (1) demographic data, including age and sex, etc.; (2) Laboratory test results, such as alanine aminotransferase (ALT), aspartate aminotransferase (AST), alpha-fetoprotein (AFP), and total bilirubin, with values selected from the most recent measurements before treatment; (3) medical history information, including chief complaints, present illness history, past medical history, and family history, obtained from admission records; (4) image reports based on the last preoperative CT, ultrasound (US), or MRI examination before treatment. Comorbidities such as ascites, varices, diabetes, and splenomegaly were also included.

#### Feature selection

First, features with missing rates >70% or variance <0.01 were removed. Remaining missing values were imputed using multiple imputation by chained equations (MICE)[6]. To reduce multicollineinearity, variables with a variance inflation factor (VIF) > 100 were excluded. Given that different surgical approaches may involve distinct predictors, we constructed multivariable logistic regression models for each procedure, using backward stepwise selection with a significance threshold of P < 0.05. Regression coefficients and their corresponding P values were provided in Supplement 1. Key predictors identified across all models were aggregated into a final set of 66 core predictive variables.

#### Outcomes

Initial treatments for HCC were categorized into four groups: laparoscopic liver resection (LLR), open liver resection (OLR), radiofrequency ablation (RFA), and TACE or chemotherapy-based treatments. Other surgical treatments were not included due to the exclusion of liver transplant recipients.

### Model development

Prior studies predicting treatments for HCC have primarily relied on structured clinical data, with limited integration of unstructured text from medical records and image reports. However, electronic health records contain critical clinical information in natural language formats, such as chief complaints, disease progression, surgical indications, and imaging descriptions, that significantly inform preoperative decision-making.

To harness semantic information from unstructured data, we employed a deep embedding model to encode medical history and image reports. Specifically, BGE-M3[7] model transformed text into vector representations, capturing latent clinical features. To mitigate high-dimensional complexity and enhance generalizability, we applied singular value decomposition (SVD) to reduce embedding dimensions to 200-dimensional vectors.

These low-dimensional embeddings were horizontally concatenated with clinical features (e.g., demographics, lab results, comorbidities) to create a hybrid feature input. Based on this framework, we developed an enhanced ensemble learning model, Embedding-Augmented Extra Trees (ET-Emb), to predict the likelihood of four primary treatment strategies.

To evaluate predictive performance, we compared ET-Emb with other machine learning algorithms widely used in treatment prediction tasks: CatBoost[8], XGBoost[9], Random Forest (RF)[10], and the multi-model voting classifier Ensemble Voting Machines (EVM)[4]. All models were internally validated using 10-fold cross-validation on the development cohort. Primary evaluation metrics included area under the receiver operating characteristic curve (ROC-AUC) and area under the precision-recall curve (PR-AUC), providing comprehensive assessment of model performance across both overall and subgroup prediction tasks. ROC-AUC for overall discriminative ability and PR-AUC, which is particularly informative for imbalanced datasets, to assess performance on the positive class.

### Model interpretability

To enhance clinical interpretability, we applied the SHapley Additive exPlanations (SHAP) framework[11] to quantify feature importance and visualize the direction and magnitude of each input variable’s contribution to treatment predictions. This analysis provides insights into how structured and unstructured features are integrated by the ET-Emb and supports its potential use in clinical settings.

### External validation

We evaluated the performance of the 10-fold cross-validated model in an external validation cohort across all treatment prediction tasks and subtasks. The same evaluation metrics used in the development phase were applied, and the mean and standard deviation of these metrics were reported as final performance estimates.

### Statistic analysis

Baseline characteristics of the internal development and external validation cohorts were summarized using descriptive statistics. Group differences were assessed using appropriate statistical tests: chi-square tests for categorical variables, and Student’s t-test or Mann– Whitney U test for continuous variables after Shapiro–Wilk normality testing.

The machine learning models were implemented in Python using PyTorch[12] version 1.12.1, scikit-learn version 1.4.2, catboost[13] version 1.2.8, and xbgoost[14] version 3.0.0 libraries. Statistical analyses were performed using SciPy and Statsmodels version 0.14.1. All computations were conducted on a GPU platform equipped with an NVIDIA RTX 3090 graphics card to ensure computational efficiency.

## Results

### Descriptive statistics

This study included 1043 development cohort patients and 55 external validation cohort patients. The two cohorts demonstrated comparable baseline characteristics (Table 1), with no significant differences in age (64.52 vs. 64.49 years, p = 0.904), gender distribution (73.86% vs. 70.49% male, p = 0.666), tumor diameter, liver function indices, or primary clinical features. The five main treatments (OLR, LLR, TACE, RFA, and Chemotherapy) were evenly distributed between cohorts (p > 0.05), demonstrating no statistically significant differences in the distribution of these treatment modalities between the two cohorts.

**Table 1.** Baseline characteristics of the development and external validation cohorts.

| Variables |  | Development cohort<br>N=1043 | External cohort<br>N=55 | p-value |
| --- | --- | --- | --- | --- |
| **Case Num.** |  | 1094 | 61 |  |
| **Age, year** |  | 64.52 (12.12) | 64.49 (11.66) | 0.904 |
| **Gender, N** |  |  |  |  |
|  | Male | 808 (73.86) | 43 (70.49) | 0.666 |
|  | Female | 286 (26.14) | 18 (29.51) |  |
| **Marital status, N** |  |  |  |  |
|  | Married | 1066 (97.44) | 57 (93.44) | 0.218 |
|  | Single | 6 (0.55) | 1 (1.64) |  |
|  | Widowed | 4 (0.37) | 0 (0.00) |  |
|  | Divorced | 2 (0.18) | 0 (0.00) |  |
|  | Other | 16 (1.46) | 3 (4.92) |  |
| **Employment status, N** |  |  |  |  |
|  | Employed | 150 (13.71) | 8 (13.11) | 0.028 |
|  | Unemployed | 181 (16.54) | 4 (6.56) |  |
|  | Retired | 346 (31.63) | 15 (24.59) |  |
|  | Other | 417 (38.12) | 34 (55.74) |  |
| **ABO blood group, N** |  |  |  |  |
|  | A | 152 (13.89) | 8 (13.11) | 0.324 |
|  | B | 140 (12.80) | 3 (4.92) |  |
|  | O | 136 (12.43) | 10 (16.39) |  |
|  | AB | 43 (3.93) | 4 (6.56) |  |
|  | Unclear | 623 (56.95) | 36 (59.02) |  |
| **LOS, days** |  | 12.24 (6.41) | 11.33 (7.17) | 0.039 |
| **Mean tumor diameter, cm** |  | 4.19 (2.52) | 4.11 (1.86) | 0.598 |
| **Mean tumor count, N** |  | 0.85 (0.63) | 0.87 (0.64) | 0.764 |
| **Inpatient expenses, Yuan** |  | 28029.41 (20150.62) | 25130.92 (19016.57) | 0.130 |
| **Comorbidities, N** |  |  |  |  |
|  | Ascites | 159 (14.53) | 9 (14.75) | 1.000 |
|  | Splenomegaly | 139 (12.71) | 10 (16.39) | 0.522 |
|  | Hypertension | 486 (44.42) | 27 (44.26) | 1.000 |
|  | Diabetes | 248 (22.67) | 13 (21.31) | 0.929 |
|  | Varix | 85 (7.77) | 6 (9.84) | 0.735 |
| **Treatments, N** |  |  |  |  |
|  | OLR | 145 (13.25) | 10 (16.39) | 0.612 |
|  | LLR | 46 (4.20) | 2 (3.28) | 0.982 |
|  | RFA | 59 (5.39) | 4 (6.56) | 0.920 |
|  | TACE | 313 (28.61) | 13 (21.31) | 0.277 |
|  | Chemotherapy | 167 (15.27) | 13 (21.31) | 0.278 |

### Model performance

The predictive performance of the ET-Emb model and benchmark models in both the development and external validation cohorts is summarized in Table 2. In the development cohort, the ET-Emb model achieved the highest overall performance in both ROC-AUC (0.84 ± 0.04) and PR-AUC (0.55 ± 0.06), compared with RF, CatBoost, XGBoost, and EVM, demonstrating superior overall predictive performance (Table 2). Similarly, in the external validation cohort, ET-Emb also achieved the highest performance, with ROC-AUC of 0.77 ± 0.02 and PR-AUC of 0.47 ± 0.03, higher than all other models. These results confirm its robust generalizability in independent datasets.

**Table 2.** Performance of Different Machine Learning Models in Development and Development cohort.

| Models | Development Cohort |  | External validation cohort |  |
| --- | --- | --- | --- | --- |
|  | ROC-AUC | PR-AUC | ROC-AUC | PR-AUC |
| Catboost | 0.80±0.03 | 0.50±0.06 | 0.74±0.01 | 0.41±0.03 |
| XGBoost | 0.78±0.04 | 0.48±0.06 | 0.73±0.02 | 0.38±0.04 |
| RF | 0.80±0.02 | 0.49±0.06 | 0.73±0.03 | 0.39±0.05 |
| EVM | 0.80±0.03 | 0.50±0.06 | 0.74±0.03 | 0.40±0.04 |
| ET-Emb | **0.84±0.04** | **0.55±0.06** | **0.77±0.02** | **0.47±0.03** |

The detailed performance metrics for each of the five treatment prediction tasks in the development cohort are presented in Table 3. In the external validation cohort, ET-Emb consistently maintained the highest performance across all five treatment tasks (Table 3, Table 4, Fig 2). While other models showed comparable metrics in some tasks (e.g., TACE, chemotherapy), ET-Emb retained superiority in chemotherapy prediction with ROC-AUC (0.78 ± 0.05) and PR-AUC (0.47 ± 0.07), exceeding CatBoost and RF.

**Fig 2.**
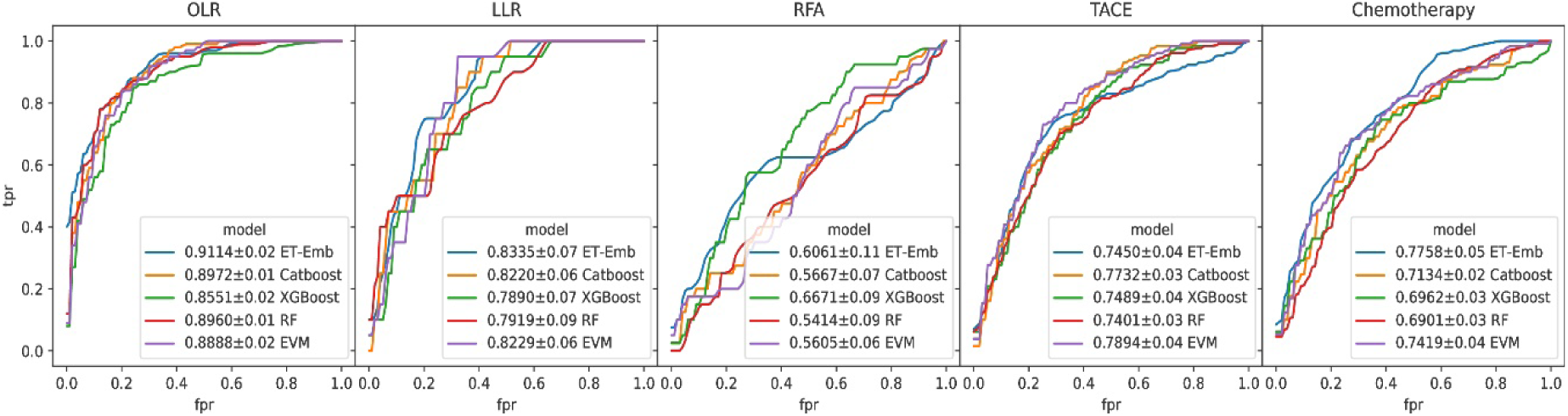
ROC-AUC on the external validation cohort for each treatment prediction task.

**Table 3.** Model performance on the development cohort for each treatment prediction task.

| Models | OLR |  | LLR |  | RFA |  | TACE |  | Chemotherapy |  |
| --- | --- | --- | --- | --- | --- | --- | --- | --- | --- | --- |
|  | ROC-AUC | PR-AUC | ROC-AUC | PR-AUC | ROC-AUC | PR-AUC | ROC-AUC | PR-AUC | ROC-AUC | PR-AUC |
| Catboost | 0.94±0.03 | 0.73±0.08 | 0.90±0.06 | 0.33±0.14 | 0.64±0.09 | 0.15±0.13 | 0.75±0.04 | 0.55±0.07 | 0.78±0.05 | 0.42±0.08 |
| Xgboost | 0.93±0.03 | **0.73±0.05** | 0.86±0.08 | 0.30±0.18 | 0.62±0.14 | 0.14±0.11 | 0.72±0.05 | 0.52±0.07 | 0.76±0.04 | 0.36±0.11 |
| RF | 0.93±0.04 | 0.70±0.08 | 0.88±0.08 | 0.32±0.16 | 0.66±0.10 | 0.14±0.10 | 0.75±0.04 | 0.54±0.06 | 0.77±0.05 | 0.39±0.10 |
| EVM | 0.94±0.04 | 0.73±0.08 | 0.89±0.08 | 0.34±0.15 | 0.66±0.10 | 0.15±0.12 | 0.75±0.04 | 0.55±0.07 | 0.78±0.04 | 0.40±0.08 |
| ET-Emb | **0.94±0.02** | 0.72±0.08 | **0.93±0.06** | **0.46±0.18** | **0.73±0.12** | **0.23±0.16** | **0.78±0.05** | **0.62±0.09** | **0.80±0.06** | **0.50±0.08** |

**Table 4.** Model performance on the external validation cohort for each treatment prediction task.

| Models | OLR |  | LLR |  | RFA |  | TACE |  | Chemotherapy |  |
| --- | --- | --- | --- | --- | --- | --- | --- | --- | --- | --- |
|  | ROC-AUC | PR-AUC | ROC-AUC | PR-AUC | ROC-AUC | PR-AUC | ROC-AUC | PR-AUC | ROC-AUC | PR-AUC |
| Catboost | 0.90±0.01 | 0.62±0.05 | 0.82±0.06 | 0.23±0.13 | 0.57±0.07 | **0.19±0.09** | 0.77±0.03 | 0.47±0.07 | 0.71±0.02 | 0.38±0.06 |
| Xgboost | 0.86±0.02 | 0.54±0.05 | 0.79±0.07 | 0.13±0.05 | **0.67±0.09** | 0.16±0.09 | 0.75±0.04 | 0.45±0.08 | 0.70±0.03 | 0.39±0.08 |
| RF | 0.90±0.03 | 0.63±0.03 | 0.79±0.09 | 0.22±0.08 | 0.54±0.09 | 0.10±0.03 | 0.74±0.03 | 0.46±0.09 | 0.69±0.01 | 0.35±0.06 |
| EVM | 0.89±0.02 | 0.60±0.04 | 0.82±0.06 | 0.19±0.08 | 0.56±0.06 | 0.17±0.10 | **0.79±0.04** | 0.47±0.10 | 0.74±0.04 | 0.39±0.07 |
| ET-Emb | **0.91±0.02** | **0.77±0.04** | **0.83±0.07** | **0.25±0.19** | 0.60±0.11 | 0.16±0.06 | 0.75±0.04 | **0.48±0.08** | **0.78±0.05** | **0.47±0.07** |

## Discussion

In this study, we successfully developed and externally validated ET-Emb, a multimodal predictive model for informing initial treatment selection in patients with HCC. The model integrates structured clinical features with rich semantic information derived from unstructured medical history and radiological reports. ET-Emb consistently outperformed established machine learning models in predicting five distinct treatment modalities (OLR, LLR, RFA,TACE, and chemotherapy) across both development and external validation cohort. Importantly, SHAP analysis revealed that the integrated text features played a substantial role in the model’s predictive power, particularly for certain subgroups like RFA.

Our findings demonstrate the effectiveness of integrating diverse data modalities for personalized HCC treatment selection, a critical area where current models often rely on limited data types[4,5]. Unlike prior studies that typically focus on a narrow range of interventions, our ET-Emb model provides intelligent recommendations across five major treatment modalities, thereby encompassing the majority of commonly employed interventions for liver cancer. This comprehensive scope more closely aligns with the complex decision-making processes observed in real-world clinical practice, where multiple viable options may exist for a given patient.

By integrating embedded representations of medical history and imaging reports, ET-Emb achieved optimal performance on both internal and external validation datasets, with average ROC-AUC values of 0.84 and 0.77, respectively. Although the prediction task for RFA was affected by the limited sample size, ET-Emb still demonstrated the best performance in this subgroup by leveraging multimodal data. Notably, SHAP analysis (Fig 3) also revealed that text features contributed substantially to model predictions— particularly for the RFA subgroup, where text features accounted for more than half of the top influential features.

**Fig 3.**
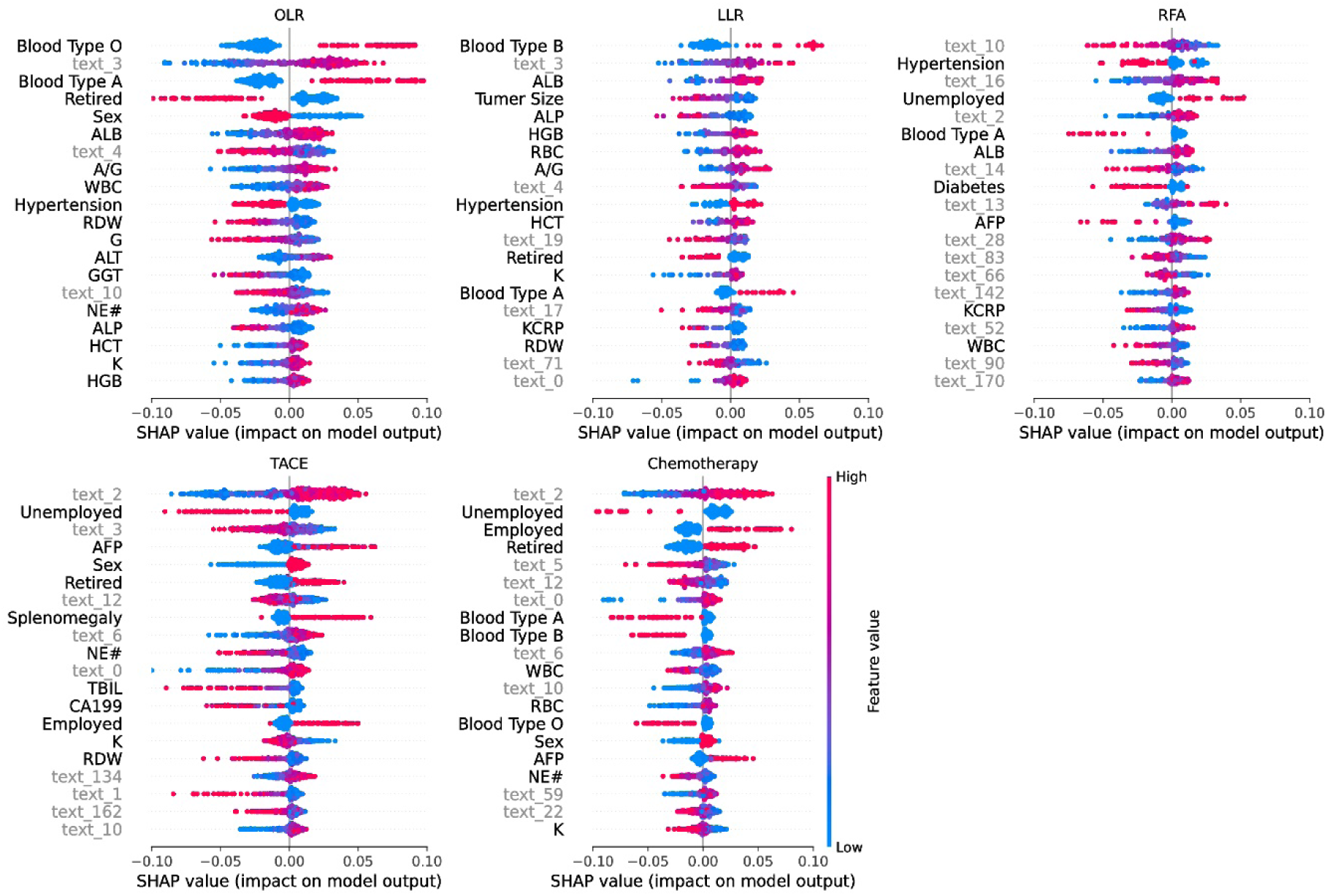
Top 20 most influential features based on SHAP across all treatment prediction tasks.

Furthermore, our model’s interpretability, facilitated by SHAP, provided insights into the clinical relevance of both structured and unstructured features. We observed that the direction of impact for many structured variables was consistent with traditional multivariable regression findings. For instance, blood types A and O were associated with an increased likelihood of liver resection, aligning with some previous research exploring the association between ABO blood types and HCC prognosis[15,16]. Similarly, the identified influence of comorbidities such as hypertension, diabetes, and splenomegaly on treatment recommendations reflects their known roles as risk factors for postoperative complications or indicators of patient fitness for specific procedures[17,18].

The inclusion and observed importance of socioeconomic factors, such as current employment status, in treatment prediction offers a novel perspective. While specific associations (e.g., male sex and retired status with chemotherapy/TACE recommendations, and their inverse association with liver resection) warrant further investigation, they suggest that factors beyond purely medical indicators may subtly influence clinical decisions or patient preferences in a real-world setting.

## Limitations

Our study has several limitations. First, we lacked long-term survival outcomes following treatment. Future work is crucial to integrate follow-up data to evaluate the model’s prognostic predictive capabilities and its impact on patient long-term benefits. Second, our model was developed and validated using data from a single institution. While the internal validation across different time periods demonstrates temporal robustness, collaboration with regional and multi-center health centers in the future will be essential to expand both the training and validation datasets, enhancing the model’s external generalizability to diverse patient populations and clinical practices. Third, we used clinical decisions made by physicians as the reference standard for model training, rather than strictly adherence to formal clinical guidelines. While this approach captures the real-world complexity of decision-making, it means the model learns from existing practices, which may encompass variations influenced by factors beyond strict medical criteria, including socioeconomic considerations. Future model development could explore how to appropriately balance these real-world influences with adherence to established guidelines. Lastly, the relatively small sample size for certain subgroups, particularly RFA, might have affected the stability of predictions for those specific modalities, although the multimodal data integration appeared to mitigate some of this effect.

## Conclusion

In conclusion, the ET-Emb model holds significant potential for clinical application in HCC treatment selection. By providing intelligent recommendations that integrate both structured and unstructured patient data, it could serve as a valuable decision-support tool for clinicians, particularly for less experienced practitioners or in complex cases. It can streamline the treatment planning process, potentially reduce variability in care, and facilitate shared decision-making with patients by clearly outlining potential treatment pathways based on their comprehensive profile.

## Data Availability

For Study Protocols: No datasets were generated or analysed during the current study. All relevant data from this study will be made available upon study completion.

## Funding

This work was supported by the Wuxi People’s Hospital Doctoral Talent Fund (Grant no. BSRC202408), the National Institute of Hospital Administration Project (Grant no. GA2024ZD02). We acknowledge the Wuxi Statistical Information Center for providing access to inpatient records and technical support during data collection.

### Competing Interests

N/A.

### Ethics approval and consent to participate

This retrospective study adhered to the Declaration of Helsinki and was approved by the Ethics Review Board of Wuxi People’s Hospital, Nanjing Medical University (KY24160). Consent to participate was waived by an Institutional Review Board in this study. The name of Institutional Review Board: the Ethics Review Board of Wuxi People’s Hospital, Nanjing Medical University.

### Data Availability

Deidentified patient data are available upon reasonable request from the corresponding author. Access to data from the Wuxi Statistical Information Center is subject to institutional and legal restrictions.

### Author Contributions

Concept and design: Wei Feng, Suyi Liu. Data acquisition: Zhiyun Yang, Yiru Tao. Statistical analysis and interpretation: Wei Feng, Xiangqian Gu. Drafting of the manuscript: Wei Feng. Critical revision of the manuscript: All authors. Supervision: Wei Jin.

## Supporting information

S1 Table Descriptive table for lab findings.

S2 Table Feature importances for OLR.

S3 Table Feature importances for LLR.

S4 Table Feature importances for RFA.

S5 Table Feature importances for TACE.

S6 Table Feature importances for Chemotherapy

## Notes

### Competing Interest Statement

The authors have declared no competing interest.

### Author Declarations

This retrospective study adhered to the Declaration of Helsinki and was approved by the Ethics Review Board of Wuxi People's Hospital, Nanjing Medical University (KY24160).

